# Plasma cell-free DNA as a prognostic biomarker in small cell lung cancer patients

**DOI:** 10.1101/2022.03.25.22272967

**Authors:** Patricia Mondelo-Macía, Jorge García-González, Alicia Abalo, Manuel Mosquera-Presedo, Rafael López-López, Luis León-Mateos, Laura Muinelo-Romay, Roberto Díaz-Peña

## Abstract

**Background:** Lack of biomarkers for treatment selection and monitoring in small-cell lung cancer (SCLC) patients with the limited therapeutic options, result in poor outcomes. Therefore, new prognostic biomarkers are needed to improve their management. The prognostic value of cell-free DNA (cfDNA) and circulating tumor cells (CTCs) have been less explored in SCLC.

**Methods:** We quantified cfDNA in 46 SCLC patients at different times during therapy. Moreover, CTCs were analyzed in 21 patients before therapy onset using CellSearch® system. Both biomarkers were associated with patients’ outcomes and a prognostic model was developed.

**Results:** High cfDNA levels before therapy were associated with shorter progression-free survival and overall survival. Furthermore, changes in cfDNA levels between baseline and 3 weeks and cfDNA levels at progression disease were also associated with patients’ outcomes. Multivariate analyses confirmed the independence of cfDNA levels as a prognostic biomarker. Finally, the three-risk category prognostic model developed included ECOG Performance Status, gender and baseline cfDNA levels was associated with a higher risk of progression and death.

**Conclusions:** We confirmed the prognostic utility of cfDNA in SCLC patients before and during therapy. Our novel risk prognostic model in clinical practice will allow to identify patients who could benefit with actual therapies.

## INTRODUCTION

Small-cell lung cancer (SCLC), which accounts for 15% of all lung cancer cases, is characterized by its aggressiveness, its strong association with tobacco and the poor outcome. About 70% of patients present extensive disease (ED-SCLC) where only 2% survive 5 years after diagnosis (1–3). For many years, chemotherapy was the unique option to treat this tumor type. However, the scenario has changed in the last years (3). New therapies, such as immunotherapy, have been recently incorporated into the management of SCLC patients and, although some survival improvements have been reported in the patients with ED-SCLC (4–8), the majority of them do not benefit from this new treatment (9). The genomic profile of SCLC is characterized by extensive chromosomal rearrangements and a high mutational burden, including in nearly all, inactivation of the tumor suppressor genes *TP53* and *RB1* (10). However, nowadays the selection of treatment in SCLC patients is not dependent on the characteristics of the tumor (11), and the criteria to stratify patients is not clear, since no predictive biomarkers have been validated for the clinical practice (12). In this context, the use of liquid biopsies as a tool to guide treatment and/or for monitoring the patients’ response represent a valuable alternative (13).

Circulating tumor DNA (ctDNA), derived from tissue tumor cells, has demonstrated its clinical utility and represents a promising tool for guiding precision medicine in several cancer types (14,15). In SCLC, different studies have investigated the importance and the clinical value of analyzing ctDNA levels. However, driver mutations known in SCLC are limited to *RB1* and *TP53* genes (16). In contrast, total cell-free DNA (cfDNA) consists of a heterogeneous and complex DNA fraction released in body fluids by any cell type through cell death mechanisms (17,18). The short half-life of cfDNA enables real-time monitoring for response or relapse, being an easy-to-implement biomarker to monitor cancer evolution and response to therapy (19).

Circulating tumor cells (CTCs) are another frequent circulating biomarker investigated in cancer. As a high metastatic tumor type, SCLC is characterized by a strong release of CTCs, with detection rates of 60.2%-94% (16), suggesting that CTCs could be employed as a disease surrogate in SCLC. The analysis of CTCs originated from the primary or metastatic sites (20) as a prognostic biomarker has been reported in different cancer types (21–23) including SCLC. However, the prognostic threshold in SCLC has been not well established (24–28). Despite their different nature, the combined analysis of total cfDNA and CTCs in patients with SCLC could provide complementary information for improving SCLC patients’ management.

In this study, we hypothesized that total cfDNA levels can serve as a useful biomarker for prognostic and follow-up of SCLC patients under first line of therapy. For this purpose, we analyzed the total cfDNA levels in a cohort of 46 patients with SCLC prior to the start of therapy, at 3 weeks after the first dose, and at the time of progression of the disease. The additional value of CTCs was investigated in our cohort in order to provide a more complete view of the disease dynamics. To our knowledge, this study is the first to examine the possible role of total cfDNA levels as a prognostic and follow-up biomarker in SCLC patients.

## METHODS

### Patients and blood sample collection

Forty-six patients with new diagnosed SCLC who received first-line treatment between June 2017 and June 2021, at the Department of Medical Oncology of Complexo Hospitalario Universitario de Santiago de Compostela were enrolled in the study. In total 111 blood samples were collected at different time points: before therapy onset (baseline) (n=46), 3 weeks after therapy start (n=40) and at the progression of the disease (n=25). A control cohort of 20 healthy individuals was also included in order to select the better assay to quantify total cfDNA. All individuals signed informed consent forms approved by Santiago de Compostela and Lugo Ethics Committee (Ref: 2017/538) prior to enrolling in the study and could withdraw their consent at any time. The study was performed in accordance with the Declaration of Helsinki.

### Clinical endpoints

Progression-free survival (PFS) was defined as the time from the date of initial treatment until the date of progression disease, death or last follow-up, whichever occurred first. Progression date was defined as the date of disease progression based on RECIST (v.1.1), or the date of clinical progression if the patient discontinued the treatment due to clinical deterioration despite not meeting criteria for RECIST progression. Overall survival (OS) was defined as the time from the date of initial treatment to the date of death or the last date of follow-up.

### Sample processing and cell free DNA isolation

Peripheral blood was obtained by direct venepuncture in CellSave tubes (Menarini, Silicon Biosystems, Bologna, Italy) and processed within 96 hours after blood collection. Plasma and cellular components were separated by centrifugation at 1,600 g for 10 minutes at room temperature. Plasma was centrifugated a second time at 5,500 g for 10 minutes at room temperature in order to remove any remaining cellular debris and aliquoted for storage at -80 °C until the time of cfDNA extraction. cfDNA was isolated from 3 mL of plasma using the QIAamp Circulating Nucleic Acid Kit (Qiagen, Hilden, Germany) using a vacuum pump, according to the manufacturer’s instructions and eluted in LoBind^®^ tubes (Eppendorf AG, Hamburg, Germany).

### Total cell free DNA quantification

CfDNA levels were quantified using two different approaches: Qubit 4 Fluorometer (ThermoFisher Scientific, Waltham, MA, USA) and quantitative PCR (qPCR) method by analyzing the telomerase reverse transcriptase (*hTERT)* single-copy gene (Thermo Fisher Scientific, Waltham, MA, USA).

2 µL of the sample were employed to quantify by the fluorometric instrument Qubit 4 using the Qubit dsDNA HS Assay Kit (ThermoFisher Scientific, Waltham, MA, USA). In the other hand, samples were quantified by a qPCR assay, described previously (29). Briefly, each qPCR reaction was carried out in a final volume of 20 µL: 10 µL of TaqMan Universal Mastermix (Thermo Fisher Scientific, Waltham, MA, USA), 1 µL of *hTERT* hydrolysis probe and 2 µL of the sample. Each sample was analysed in duplicate. In addition, each plate included a calibration curve and negative controls. The calibration curve calculated based on a dilution series of a commercial standard human genomic DNA (Roche Diagnostics, Mannheim, Germany), was fragmented in 184 bp using Covaris^®^ E220 Focused-ultrasonicator (Covaris, Massachusetts, USA) using the following protocol: 430s duration, peak incident power of 175 Watts, duty factor of 10% and 200 cycles per burst. Fragments size were then determined using a TapeStation 4700 (Agilent, Santa Clara, CA, USA) and the High Sensitivity DNA ScreenTape^®^ (Agilent, Santa Clara, CA, USA). Amplification was performed under the following cycling conditions using a QuantStudio™ 3 real-time PCR system (Thermo Fisher Scientific, Waltham, MA, USA): 50 °C for 2 min; 95 °C for 10 min; 40 cycles of 95 °C 15 s; and 60 °C for 1 min. Data were analyzed with QuantStudio™ Design & Analysis software, version 2.5.1 (Thermo Fisher Scientific, Waltham, MA, USA). The final concentration of each sample was calculated by interpolation of the mean of cycle quantification values (Cq) with the calibration curve. Values with a Cq confidence interval less than 0.95 were discarded. Moreover, only assays with R^2^ values greater than 0.98 for the standard curve and with an efficiency ≥ 88.8% were used. Results obtained from both approaches (Qubit vs *hTERT* qPCR) were compared.

### CTC detection and enumeration

Circulating tumor cell analyses were performed using the CellSearch® system (Menarini, Silicon Biosystems, Bologna, Italy). Peripheral whole blood of each patient was collected in CellSave preservative tubes (Menarini, Silicon Biosystems, Bologna, Italy), stored at room temperature and processed within 96 hours after the blood was drawn.

Briefly, 7.5mL of whole blood were mixed with 6mL of buffer and centrifugated at 800 g for 10 minutes at room temperature. Next, samples were processed in the CellTracks Autoprep system using the Circulating Tumor Cell Kit (Menarini, Silicon Biosystems, Bologna, Italy). The kit consists of ferrofluids coated with epithelial cell-specific anti-EpCAM antibodies to immunomagnetically enrich epithelial cells; a mixture of antibodies directed to cytokeratins (CKs) 8, 18, and 19 conjugated to phycoerythrin (PE); an antibody to CD45 conjugated to allophycocyanin (APC); nuclear dye 4′,6-diamidino-2-phenylindole (DAPI) to fluorescently label the cells as well as buffers to fix, permeabilize, wash and resuspend the cells. Finally, samples were analyzed with the CellTracks Analyzer II according to the manufacturer’s instructions. The CTCs were identified as round or oval cells with an intact nucleus (DAPI^+^), CK positive and CD45 negative.

### Statistical analysis

Continuous data were summarized as mean, median and range whereas frequency and percentage were presented for categorical variables. Categorical variables were compared using the chi-square test or Fisher’s exact test. Swimmer plot was provided to visualize the times of sample collection, every patient’s therapy and clinical outcomes. Pearson test was used to evaluate a pairwise correlation between the different strategies to quantify the cfDNA, by fluorometry and qPCR. The Mann–Whitney–Wilcoxon U-Test was used to compare continuous variables between groups. Receiver operating characteristics (ROC) curves were computed based on cfDNA levels of SCLC patients and healthy controls, representing the area under the curve (AUC) values and computing the confidence intervals (CI) at 95% confidence levels. ROC curves were also constructed to evaluate the thresholds of baseline cfDNA levels for PFS and OS analyses. Kaplan-Meier method was used to plot the survival curves applying the log-rank test. Univariate and multivariate Cox regression analyses were used to evaluate factors independently associated with PFS and OS. A final prognostic model for PFS and OS was developed. Comparisons of Cox proportional hazard regression models were made using the Akaike information criterion (AIC) technique (30), with a smaller AIC value indicating the better model. All statistical analyses were performed using R version 4.1.1. The following R packages were used: survival (31), survminer, ggplot2 (32), pROC (33), gtsummary (34), swimplot, stats, rstatix.

## RESULTS

### Patient characteristics and sample collection

Forty-six SCLC patients were included in the study. Their clinicopathological characteristics are presented in Table 1. The median age was 67 (range 47-83) years, all the patients were current or former smokers, and most were males (84.78%) and stage IV tumors (89.13%). The Eastern Cooperative Oncology Group Performance Status (ECOG PS) <2 accounted for 67.39% of cases and 52.17% of patients had a number of metastases ≥2. The median number of chemotherapy or chemotherapy/immunotherapy treatment cycles was 5 (range 1–11). At the time of analysis, 41 of the 46 (89.13%) evaluable patients had experienced disease progression and 38 of the 46 (82.6%) evaluable patients had died. Sample collection was performed before therapy onset, 3 weeks after initiation of therapy and at the time of progression disease (Figure 1). Median PFS and OS were 174 (range 4-483) and 229 (range 4-748) days, respectively.

**Table 1.**
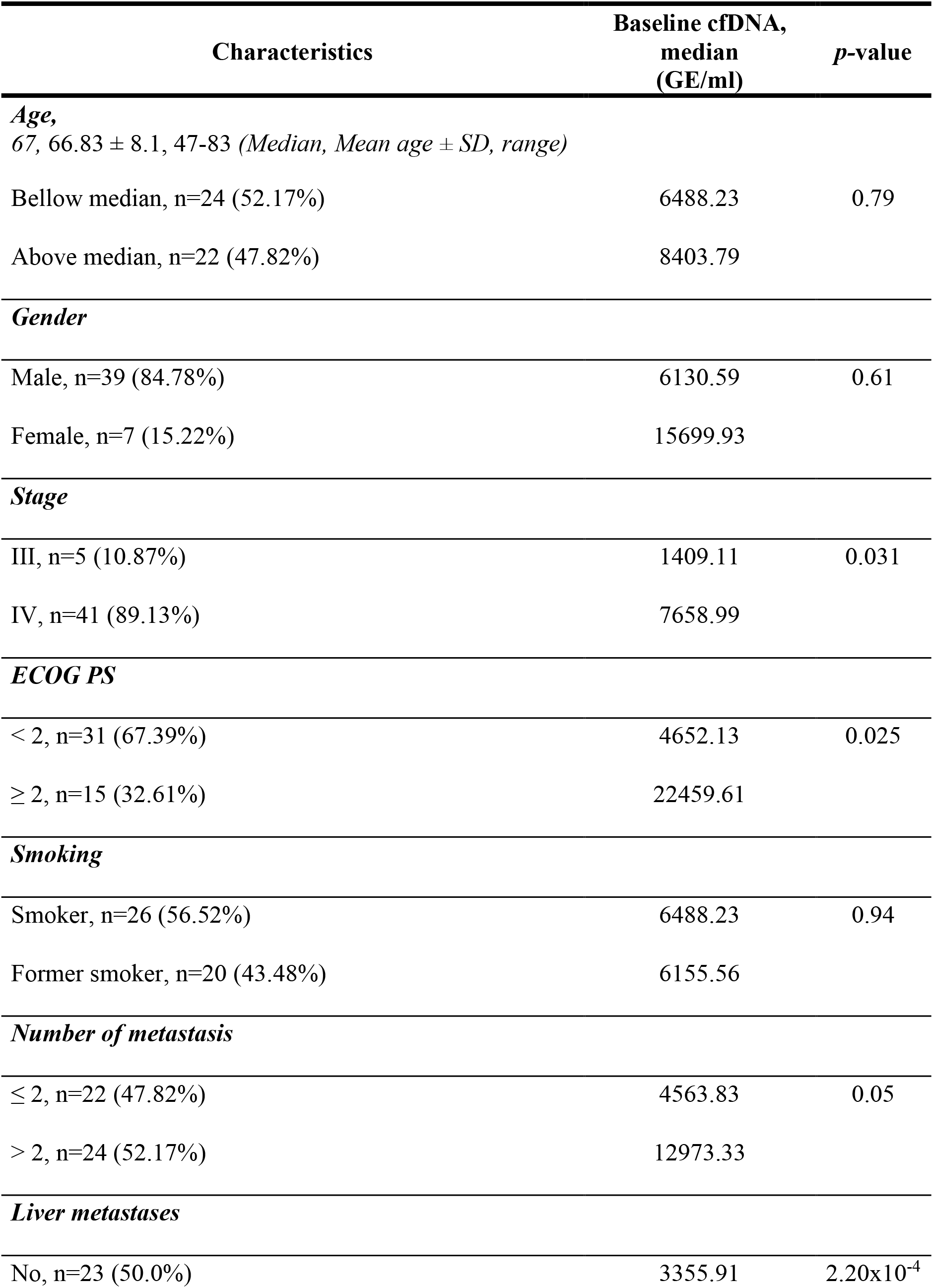
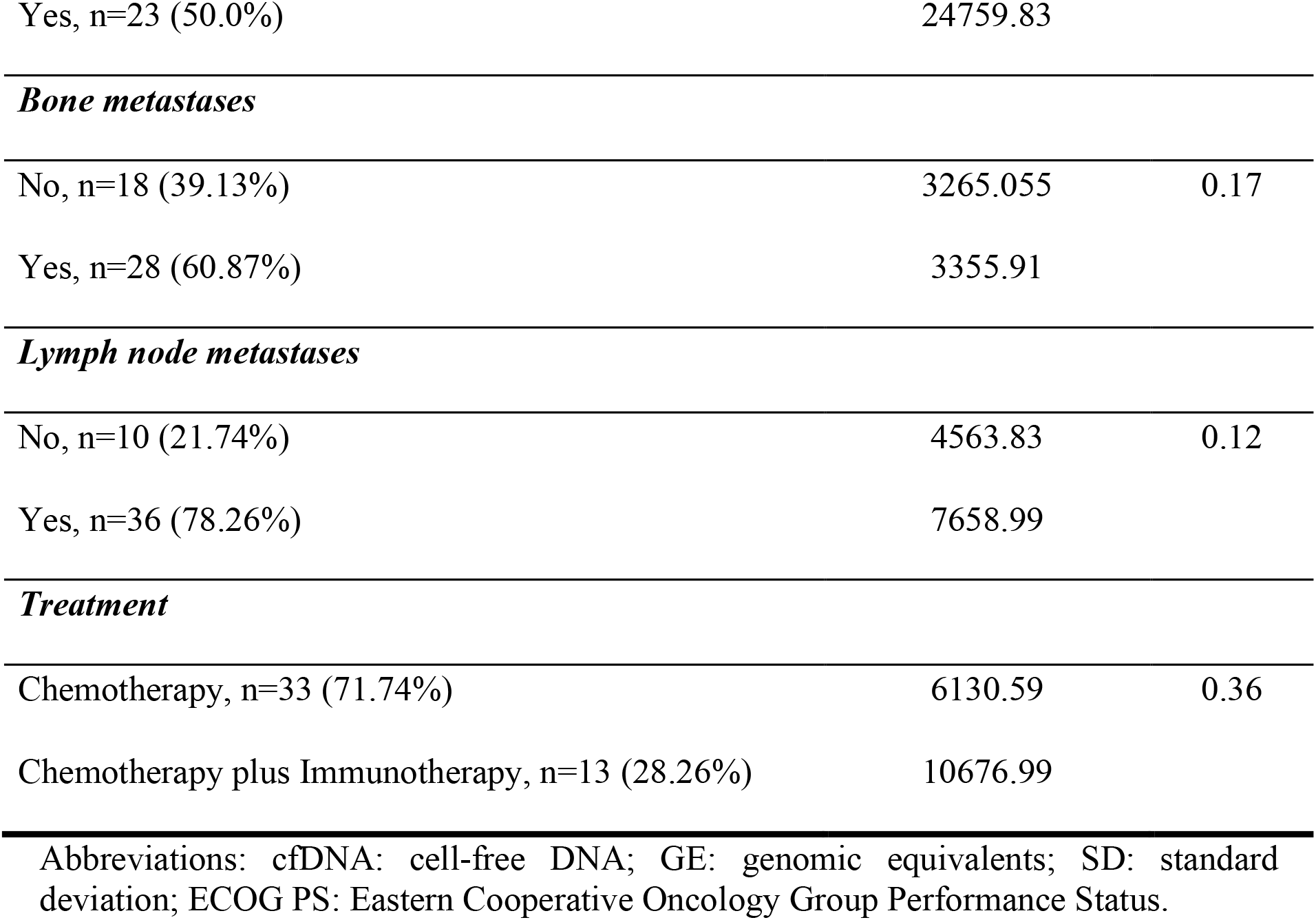
Association between patients’ demographics/clinical characteristics and baseline cfDNA levels.

**Figure 1.**
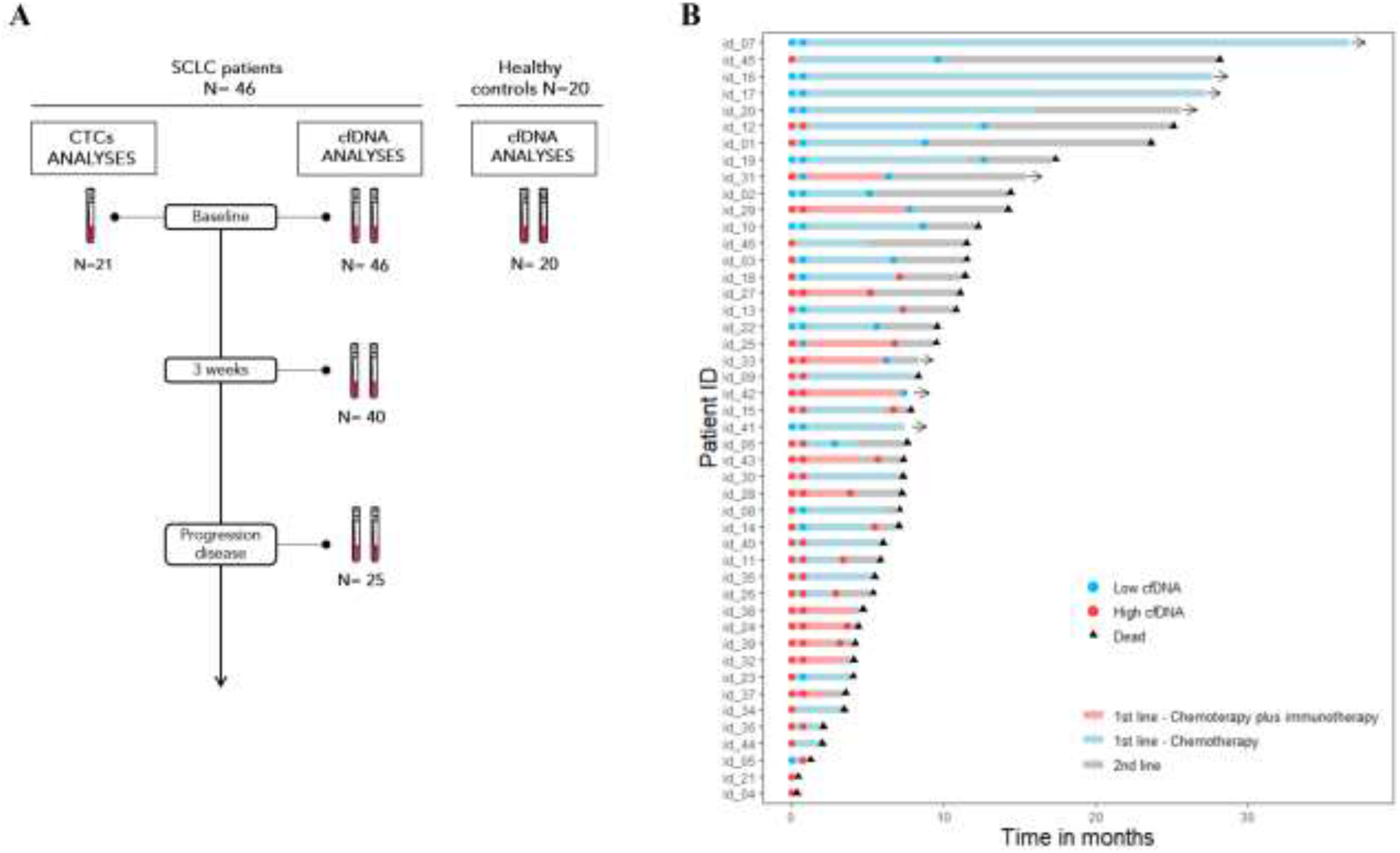
Study schema and clinical course of all patients included in the study. (A) Study sampling points and cohorts. (B) Swimmers’ plot showing each patient therapy and the different times of sample collection. The total length of each bar indicates the duration of survival from the diagnoses. Abbreviations: SCLC, small-cell lung cancer; CTCs, circulating tumor cells; cfDNA: cell free DNA.

### CfDNA levels are specifically increased in SCLC patients

In order to determine the better method to quantify the total cfDNA in patients with SCLC, we compared the Qubit 4 Fluorometer (ThermoFisher Scientific, Waltham, MA, USA) versus the qPCR method by analyzing the *hTERT* single-copy gene (Thermo Fisher Scientific, Waltham, MA, USA). A good correlation between both approaches at different times of therapy (baseline, 3 weeks, and progression disease) was found (R^2^=0.959) (Figure 2A). In addition, 20 healthy controls were included in order to compare their cfDNA levels with our patient cohort using both approaches. Total cfDNA levels in healthy controls were statistically lower than in those found in the SCLC cohort (Wilcoxon test p=1.2×10^−08^ and p=1.5×10^−11^, using both fluorometer method and qPCR assay, respectively) (Figure 2B-C). Our qPCR assay showed an AUC=0.95 (specificity 95% and sensibility 85%) whereas the fluorometer method presented an AUC=0.94 (specificity 100% and sensibility 80%) (Figure 2D-E). These results evidenced that cfDNA levels were increased as a result of the malignant disease in SCLC patients and reinforced their interest as a potential biomarker to follow-up the disease evolution. In addition, the PCR-based strategy was prioritized to quantify the cfDNA since this method appears as a more robust option than fluorometric quantification.

**Figure 2.**
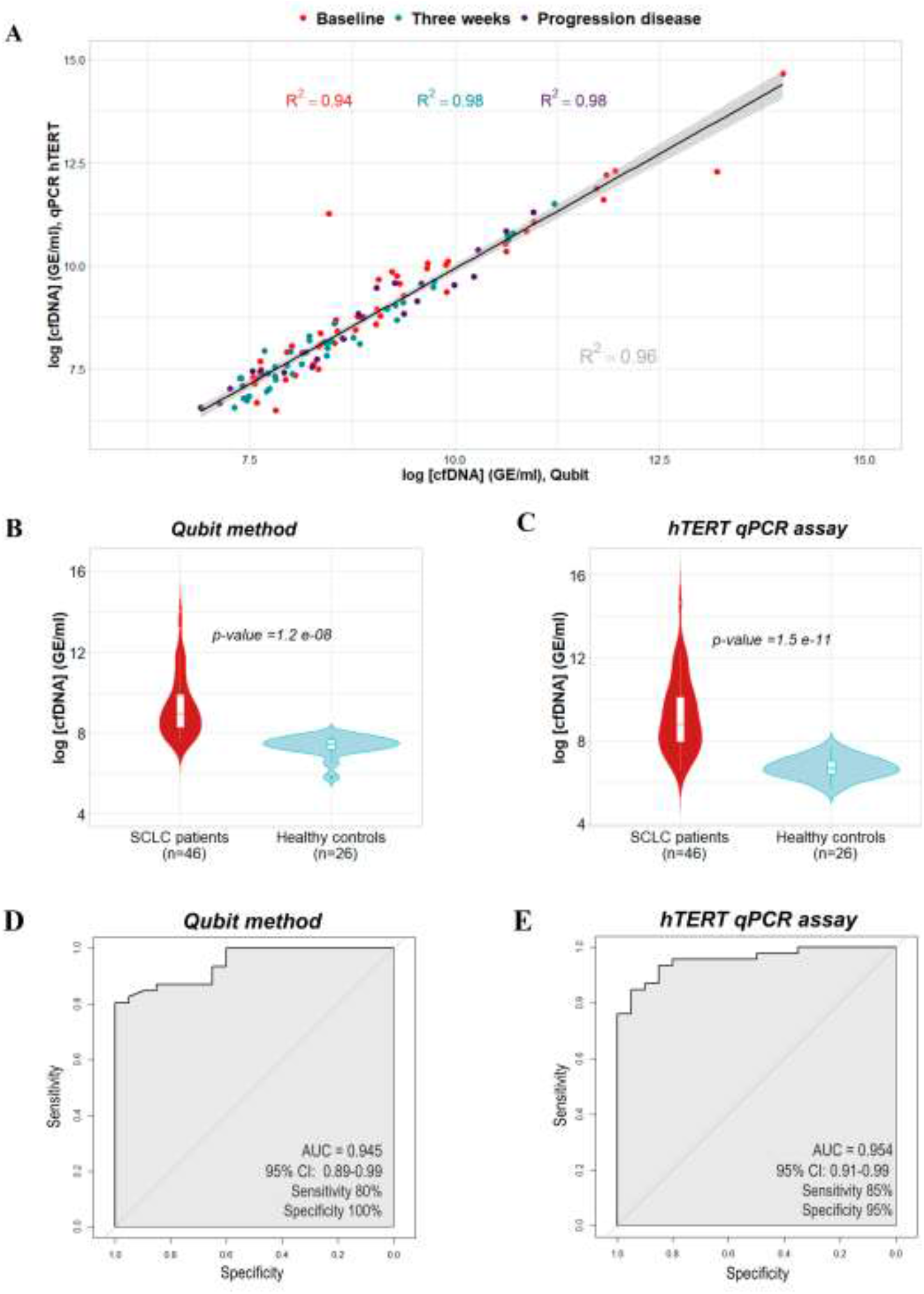
(A) Correlation between cfDNA levels using Qubit method and qPCR assay at different times of therapy (baseline, 3 weeks and progression disease). (B-C) Total cfDNA levels in healthy controls and patients with SCLC. (D-E) ROC curves for Qubit method (C) and qPCR assay (E) show high sensitivity and specificity to discriminate healthy controls and SCLC patients. Abbreviations: cfDNA, cell free DNA; GE, genomic equivalents; AUC, area under the curve; CI, confidence interval.

### Clinical interest of cfDNA analysis at baseline

Total cfDNA was quantified by qPCR assay at different time points in our patient cohort with the goal to evaluate its potential as a monitoring tool (Supplementary Table 1). CfDNA levels at baseline were significantly higher in patients with stage IV cancer, poor performance status, an elevated number of sites of metastasis and presence of liver metastases (P value≤ 0.05) (Table 1). There was no statistically significant association with respect to age, gender, smoking status, presence of bone metastasis, presence of lymph node metastases and the treatment used.

In addition, the possible role of cfDNA as a prognostic biomarker before therapy in SCLC was investigated. Thus, the cfDNA levels were log-transformed and the patients were dichotomized into high and low cfDNA level groups based on ROC analysis (Supplementary Table 2). The thresholds of baseline log cfDNA levels were chosen at 7.650 (∼2100.65 copies in hG/mL plasma) and 8.077 (∼3219.56 copies in hG/mL plasma) for PFS and OS analyses, respectively. We found that patients with high levels of total cfDNA at baseline presented shorter PFS (long rank *p=*0.0005, hazard ratio, 5.06; 95% CI 1.89–13.6) and OS (long rank *p=*0.0005, hazard ratio, 3.32; 95% CI 1.50-7.37) than those with low levels of total cfDNA (Figure 3A-B). The median PFS of patients in the low baseline cfDNA group was 350 days versus 156 days in the high baseline cfDNA group, whereas the median OS was 426 and 210 days in the two respective groups (low vs high baseline total cfDNA levels). Univariate and multivariate Cox regression analyses of PFS and OS were performed considering various clinical and demographic variables (ECOG PS, sex, age, stage, number of metastases, presence of liver metastasis and smoking status) (Table 2). Multivariate regression analyses confirmed the value of the cfDNA levels at baseline as an early independent predictor biomarker for PFS and OS (hazard ratio, 46.0; 95% CI, 3.16-672; *p*-value =0.005 and hazard ratio, 32.4; 95% CI, 3.05-344; *p*-value =0.004, respectively).

**Figure 3.**
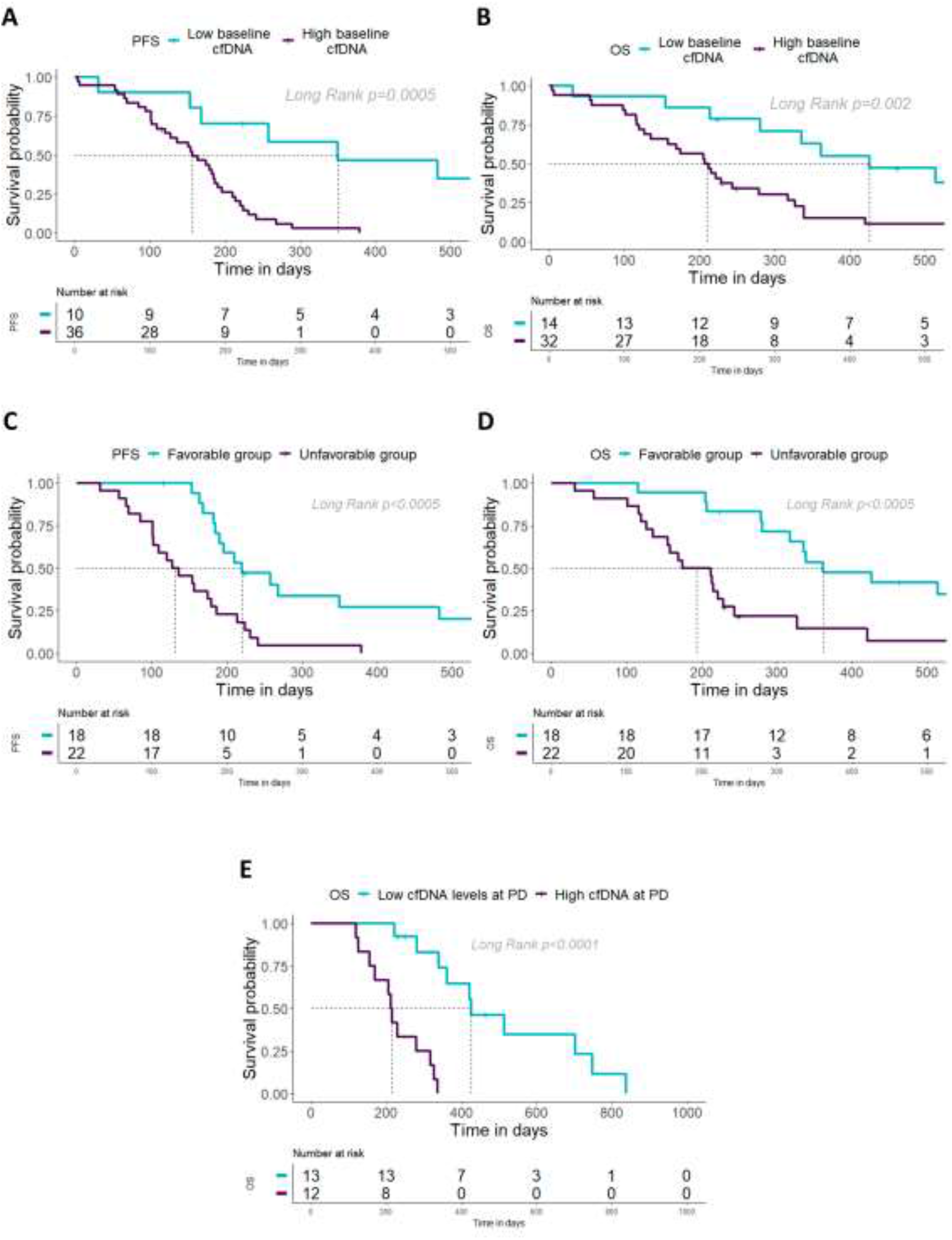
CfDNA levels as a prognostic biomarker at different time points of therapy. (A-B) Kaplan-Meier survival analysis of cfDNA levels at baseline for PFS (A) and OS (B). (C-D) Kaplan-Meier survival analysis of cfDNA levels at 3 weeks for PFS (C) and OS (D). (E) Kaplan-Meier survival analysis of cfDNA levels at progression disease for OS. Abbreviations: cfDNA, cell free DNA; OS, overall survival; PFS, progression-free survival; PD: progression disease.

**Table 2.** Univariate and multivariate Cox regression analyses of cfDNA levels, CTC counts and clinical parameters.

| Variable | Univariate |  | Multivariate |  |
| --- | --- | --- | --- | --- |
|  | p-value | HR<br>(95% CI) | p-value | HR<br>(95% CI) |
| **PFS** |  |  |  |  |
| Baseline log cfDNA (high vs. low cfDNA) | 0.001 | 5.06 (1.89-13.6) | 0.005 | 46.0 (3.16-672) |
| Baseline CTC count, CellSearch ($\geq 150$ vs. $<150$) | 0.028 | 3.47 (1.14-10.6) | | |
| ECOG ($\geq 2$ vs. $<2$) | $<0.001$ | 3.57 (1.72-7.40) | 0.04 | 17.9 (1.11-289) |
| Sex (male vs. female) | 0.2 | 1.76 (0.74-4.22) |  |  |
| Age (continue) | 0.5 | 1.01 (0.97-1.05) |  |  |
| Stage (IV vs. III) | 0.07 | 3.00 (0.91-9.91) |  |  |
| Number of metastasis ($>2$ vs. $\leq 2$) | 0.1 | 1.70 (0.90-3.19) | | |
| Liver metastasis (yes vs. no) | 0.08 | 1.74 (0.93-3.23) |  |  |
| Smoking (smoker vs. former smoker) | 0.8 | 0.92 (0.49-1.70) |  |  |
| 3 weeks log cfDNA (high vs. low cfDNA)* | $<0.0001$ | 3.50 (1.69-7.23) | 0.004 | 3.49 (1.50-8.12) |
| **OS** |  |  |  |  |
| Baseline log cfDNA (high vs. low cfDNA) | 0.003 | 3.32 (1.50-7.37) | 0.004 | 32.4 (3.05-344) |
| Baseline CTC count, CellSearch ($\geq 150$ vs. $<150$) | 0.07 | 2.71 (0.93-7.88) | | |
| ECOG ($\geq 2$ vs. $<2$) | $<0.001$ | 4.54 (2.13-9.68) | | |
| Sex (male vs. female) | 0.2 | 1.80 (0.75-4.36) |  |  |
| Age (continue) | 0.5 | 1.01 (0.97-1.06) |  |  |
| Stage (IV vs. III) | 0.08 | 2.86 (0.86-9.47) |  |  |
| Number of metastasis (>2 vs. ≤2) | 0.5 | 1.23 (0.65-2.33) |  |  |
| Liver metastasis (yes vs. no) | 0.3 | 1.45 (0.77-2.76) |  |  |
| Smoking (smoker vs. former smoker) | 0.5 | 0.82 (0.43-1.55) |  |  |
| 3 weeks log cfDNA (high vs. low cfDNA)* | <0.001 | 3.67 (1.72-7.82) | 0.002 | 4.35 (1.68-11.3) |
| PD log cfDNA (high vs. low cfDNA)* | <0.001 | 15.2 (3.28-70.7) | 0.001 | 19.5 (3.30-115) |

\*Multivariate Cox regression model including sex, age, Eastern Cooperative Oncology Group Performance Score, stage, number of metastases, presence of liver metastasis and smoking status. The levels of cfDNA were determined as low (<cut-off) or high (≥cut-off) based on the cut-off obtained from the ROC curve analyses. Abbreviations: cfDNA, cell free DNA; CTC, circulating tumor cell; ECOG, Eastern Cooperative Oncology Group Performance Score; PFS, progression-free survival; OS, overall survival; HR, hazard ratio; CI, confidence interval.

### Longitudinal analysis of total cfDNA levels during therapy

To determine whether cfDNA levels can be employed to monitor patients’ evolution during therapy, we quantified longitudinal cfDNA levels at 3 weeks after initiation of treatment (n=40) and at progression disease (n=25). We found that levels of total cfDNA were significantly higher before therapy than at 3 weeks after therapy onset (Wilcoxon test *p=* 0.002; Supplementary Figure 1), suggesting clearance of ctDNA after therapy start which impact on cfDNA levels. However, no significant differences between cfDNA levels at baseline and at progression disease were found (Supplementary Figure 1).

To analyze the possible prognostic role of cfDNA monitoring during therapy, cfDNA levels were also log-transformed and the patients were dichotomized into high and low cfDNA level groups based on ROC analysis (Supplementary Table 2) in each sample time. Thus, the possible prognostic role of monitoring cfDNA levels at 3 weeks after initiation of treatment was investigated. We divided our cohort into groups after considering their changes in cfDNA levels: Favorable group (patients with low cfDNA levels at baseline and at 3 weeks + patients with high cfDNA levels at baseline but low levels at 3 weeks; n=18) and unfavorable group (patients with low cfDNA levels at baseline but high cfDNA levels at 3 weeks + patients with high cfDNA at baseline and at 3 weeks; n=22). Patients for the unfavorable group showed a shorter PFS (long rank *p<*0.0005, hazard ratio, 3.5; 95% CI 1.69-7.23) and OS (long rank *p*<0.0005 hazard ratio, 3.67; 95% CI 1.72-7.82) (Figure 3C-D) than the favorable group. Multivariate regression analyses confirm the independence of the cfDNA changes between baseline and 3 weeks as a prognostic biomarker of PFS and OS (hazard ratio, 3.49.0; 95% CI, 1.50-8.12; *p*-value =0.004 and hazard ratio, 4.35; 95% CI, 1.68-11.3; *p*-value =0.002, respectively) (Table 2).

Finally, cfDNA quantification at the time of progression disease was performed in 25 SCLC patients. Our results reported that patients with high cfDNA levels at this time point survive fewer days than patients with low cfDNA levels (long rank *p<*0.0001, hazard ratio, 15.2; 95% CI 3.28-70.7; 426 days in the high cfDNA levels group versus 214 days in the low cfDNA levels group) (Figure 3E).

### Circulating tumor cells analyses and prognostic value

CTCs analyses were performed in 21 patients with SCLC before starting the treatment. 85.71% of patients (18 of 21 patients) presented at least 1 CTC with a median of 26 CTCs (range 0 – 4796) (Supplementary Figure 2A-B). CTCs number at baseline was significantly higher in patients with extensive disease (stage IV) and with poor performance status (Supplementary Figure 3A-J). Also, the presence of CTCs and high cfDNA levels were significantly associated (Supplementary Figure 3K), indicating that both markers are reflecting the tumor burden.

In another hand, different cut-offs were analyzed in order to determine the possible prognostic value of CTCs (Supplementary Table 3). We found that the presence of ≥150 CTCs/7.5mL of blood was significantly associated with shorter PFS rates (long rank *p=0*.*02*, hazard ratio, 4.66; 95% CI 1.11-19.6) in our cohort (Supplementary Table 3; Supplementary Figure 2C). In multivariate analysis CTCs did not show value as an independent predictive biomarker of PFS and OS.

### Prognostic model for PFS and OS

An independent prognostic model for both PFS and OS was developed. Three variables were retained in the final prognostic model: cfDNA levels (high vs. low levels), ECOG PS (<2 vs. ≥2) and sex (male vs. female). The detailed results of the multivariate analyses are shown in Figure 4A. Subsequently, we segregated patients into three risk categories: patients with all adverse prognostic factors were classified in the poor-risk category (high cfDNA levels, ECOG PS≥2 and male gender), patients with two adverse prognostic factors were classified in the intermediate-risk category, and patients with one or none adverse prognostic factor were classified in the favorable-risk category. The Kaplan–Meier curves representing the three risk categories and median PFS and OS are presented in Figure 4B-C. Median PFS ranged from 124 to 289 days based on the number of adverse prognostic factors present before therapy. Median OS ranged from 115 to 514 days.

**Figure 4.**
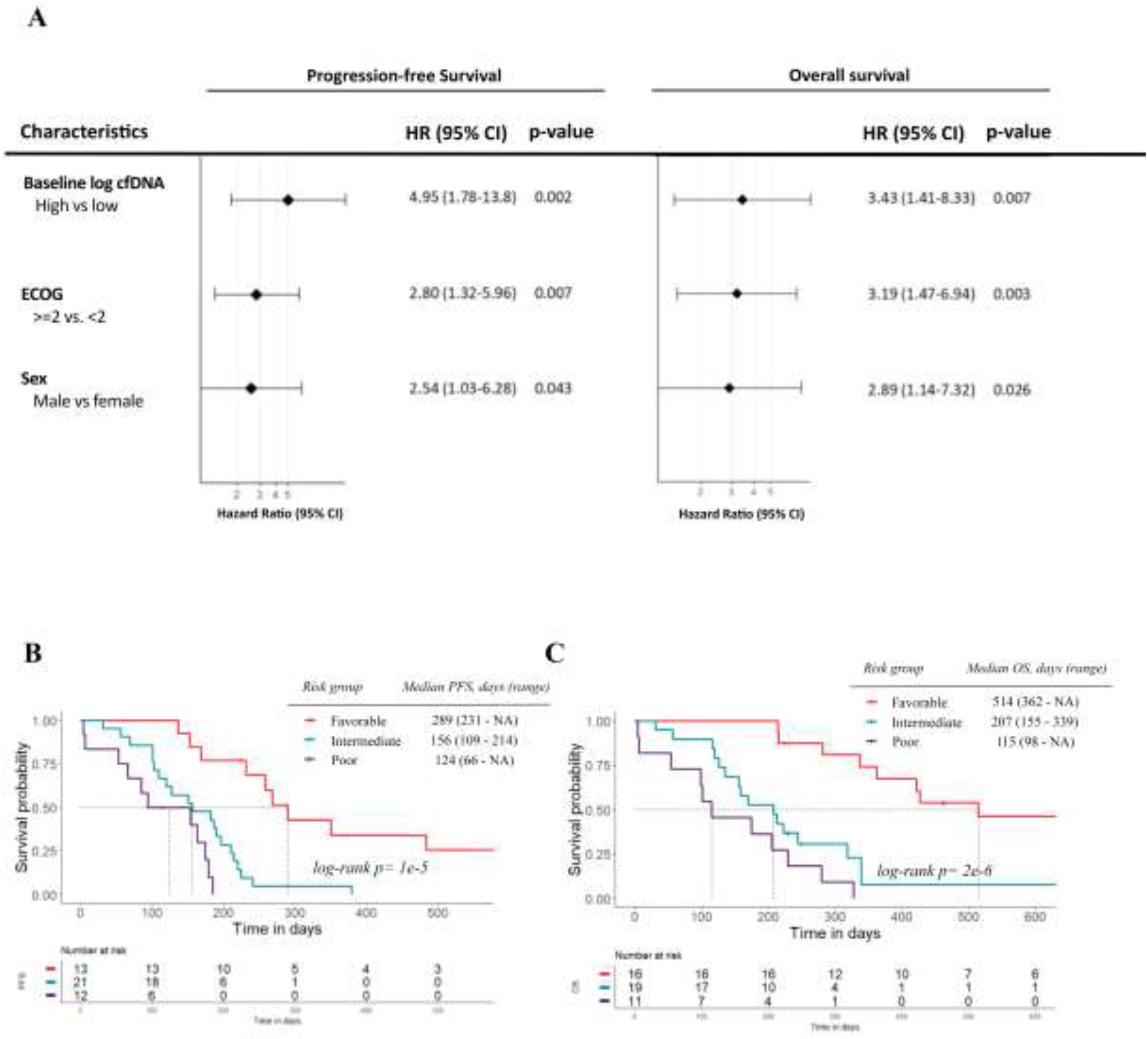
(A) Final multivariate Cox regression prognostic model for PFS and OS. (B-C) Kaplan-Meier survival analysis according to risk-group for PFS and OS. Abbreviations: cfDNA, cell free DNA; ECOG, Eastern Cooperative Oncology Group Performance Score, PFS, progression-free survival; OS, overall survival; HR, hazard ratio; CI, confidence interval.

Going up from a lower category (favorable) to upper categories (intermediate and poor), the progression model quintuples the risk of disease recurrence (HR= 5.37, 95% CI, 2.32-12,4; p-value= 3×10^−5^) and six folds the risk of death (HR= 6.02, 95% CI, 2.66-13.6; p-value= 3×10^−6^) (Supplementary Table 4).

## DISCUSSION

Precision medicine has the objective of optimizing the selection of the best therapy for each patient. In this context, liquid biopsy has emerged as a promising and minimally invasive tool for this due to its ability to provide a total image of primary and metastatic tumors at different times across therapy (35). Recently, the management of SCLC has changed and new therapies, such as immunotherapy among others, are been investigated and approved for clinical use (16,36,37). Nevertheless, the necessity to find a prognostic biomarker for helping to select the therapy prescribed and to monitor the evolution of the disease during the treatment, remains a challenge in SCLC patients. In this study, we report for the first time the possibility to employ the cfDNA and its quantification as a prognostic biomarker in SCLC prior to starting therapy and at different time points. Our analyses allow us to identify a group of low-risk patients characterized by low cfDNA levels at baseline who probably will benefit from both: chemotherapy in monotherapy or the combination of immunotherapy and chemotherapy. The study of another common circulating biomarker, CTCs, also provided us information for the prognostic of patients before starting therapy, although the results were less clear. We found a significant association between presence of a high number of CTCs (≥150 CTCs) and worse PFS. Total cfDNA refers to a heterogeneous and complex DNA fraction free released in body fluids by any cell type (not only tumoral) through several cell death mechanisms such as secretion, apoptosis and necrosis (17,18). In line with our results, it’s well reported that cancer patients present higher cfDNA levels than healthy controls (38,39) but few studies have investigated the possible prognostic and predictive value of total cfDNA quantification in patients with SCLC. In contrast, the ctDNA, the tumor-derived fraction of this cfDNA, has been reported as a prognostic and predictive biomarker in several works (40–44). Almodovar et al. reported that changes in the mutant allele frequencies on ctDNA were associated with response to treatment and relapse. Twenty-seven patients with SCLC were analysed by next-generation sequencing (NGS) custom panel, however, the lack of driver mutations known in SCLC, limited the number of genes analyzed (40). In another work, Devarakonda et al analyzed 564 patients using a larger NGS panel, including 73 genes, and, according to previous results into the bibliography, *RB1* and *TP53* were the most frequent mutant, however, any prognostic or predictive value was reported in this study.

In this way, total cfDNA quantification allows to detect the total DNA released from normal and also tumor cells into the blood. Thus, despite the few known driver mutations found in SCLC, cfDNA quantification allows to quantify the total levels before treatment and monitor the changes during therapy. Recently, our group demonstrated the feasibility to quantify cfDNA levels in non-small cell lung cancer patients and its association with patients’ outcomes (45), suggesting its possible utility in SCLC.

Thus, in the present work, we quantified total cfDNA levels using two different technologies, a fluorometric assay Qubit and a more specific assay, the qPCR assay analyzing the *hTERT* gene. CfDNA levels quantified by both technologies showed good concordance. Furthermore, the concordance of cfDNA levels at any time point of therapy using both methods also showed a good concordance. Therefore, both methods could be used to robustly measure the cfDNA content. However, to complete our study we chose the qPCR assay, which is a high sensitive and specific assay for cfDNA quantification in SCLC patients, and was previously employed in studies focused on non-small cell lung cancer (45–48).

Regarding the clinical meaning of cfDNA content, we found that high levels were significantly associated with shorter PFS and OS before therapy onset, being a robust independent prognostic biomarker in newly diagnosed SCLC patients. Also, cfDNA levels at baseline were higher in patients with stage IV that could be a consequence of more aggressive disease. This can be partially explained by an increase of ctDNA levels released from the tumor cells to the bloodstream, increasing the total cfDNA fraction. Moreover, analyses showed that changes of cfDNA levels between baseline and 3 weeks are associated with patients’ outcomes, being those patients with high values in both sample points, the ones with the worst prognosis. In addition, high cfDNA levels at the time of disease recurrence were associated with a higher risk of death. Of note, multivariate analyses showed the independence of cfDNA levels at 3 weeks and at progression disease as a prognostic biomarker. These results suggest that cfDNA monitoring could provide valuable information for the management of SCLC as our group previously reported in non-small cell lung cancer (NSCLC) (29). Thus, in clinical practice, in those SCLC patients with high levels of cfDNA at the time of disease recurrence, the selection of a more aggressive therapy or the intensification of clinical visits would be considered.

Besides cfDNA levels, we investigated the prognostic value of additional biomarkers such as CTCs and clinical characteristics. CTCs were analyzed in a cohort of 21 SCLC patients using the CellSearch® system, the only Food and Drug Administration (FDA)-approved device for CTC enumeration in prostate, breast and colorectal cancer. According to previous studies (16), a detection rate of 85.71% was found in our study. Moreover, the CTC count at baseline determined using the CellSearch® system was significantly associated with PFS and OS (24,27,28,49,50). For example, Naito et al. reported that the presence of ≥8 CTCs/7.5mL of blood was associated with worse OS (24), however, another study employed 50 CTCs as cut-off for PFS and OS (28). In fact, a consensus regarding the optimal cut-off of CTCs and the prognostic value remains a challenge (16). In this work, we found a discrete association between the presence of ≥150 CTCs and a shorter PFS, however multivariate analyses did not show independent value for the CTC count. Interestingly, high cfDNA levels and the presence of CTCs at baseline were significantly associated, reporting the clear association between both circulating biomarkers. CTCs release in the bloodstream is related to the intravasation process of potentially metastatic cancer cells. Nevertheless, cfDNA is released by any cell type including tumoral and normal cells, however, how cfDNA release relates to tumor biology is currently unknown. Finally, we evaluate several factors that could influence the patients’ outcomes. Thus, we proposed a simple model to segregate patients into three categories based on risk of progression and death (taking into account the cfDNA levels, ECOG and gender of patients). We found that patients with one or less adverse prognostic factors were classified in the favorable-risk category and present a longer PFS and OS. Thinking about the clinical relevance of these results, some limitations in our design should be considered. First, the sample size of our CTC cohort was relatively small and CTC monitoring during therapy could provide more valuable information. Finally, a validation study of our prognostic model in a larger cohort of patients can arise more robust conclusions.

In conclusion, we describe an important potential role of cfDNA levels as a prognostic biomarker in newly diagnosed SCLC patients and also could provide useful information about disease evolution. In addition, a prognostic model employing cfDNA levels and some clinical characteristics (ECOG and gender) allow us to stratify patients and detect those who particularly could benefit from treatment.

## Data Availability

The data that support the findings of this study are available from the corresponding author upon reasonable request.

## Acknowledgments

This project never could be possible without the kindly collaboration of all patients.

## Author’s contributions

All authors agreed to submit this manuscript. L.L-M., L.M-R., R.L-L and R.D-P. designed and supervised the project. M.M-P., J.G-G. and L.L-M. contributed clinical samples and patient characteristics. P.M-M and A.A performed experiments. P.M-M. and R.D-P analyzed the data. P.M-M. and R.D-P prepared the manuscript. L.M-R. and L.L-M supervised the manuscript. All the authors read and approved the final manuscript.

## Ethics approval and consent to participate

The present study was approved by the ethic committee of Santiago de Compostela and Lugo Ethics Committee (Ref: 2017/538). Written informed content was obtained from every participant prior to enrolling in the study and could withdraw their consent at any time. The study was performed in accordance with the Declaration of Helsinki.

## Consent for publication

Not applicable.

