## Supplemental material for "Plasma cell-free DNA as a prognostic biomarker in small cell lung cancer patients"

### SUPPLEMENTARY TABLES

**Supplementary table 1.** Statistics of cfDNA levels at baseline, at 3 weeks after the onset of therapy and at disease progression.

| | **Sample size** | **Mean (GE/mL)** | **Median (GE/mL)** | **Standar desviation** | **Range (GE/mL)** |
|---|---|---|---|---|---|
| Baseline, cfDNA levels | 46 | 81174.03 | 6488.23 | 342126.8 | 660.7-2320370.4 |
| Three weeks post-treatment, cfDNA levels | 40 | 9034.88 | 2912.55 | 18500.36 | 709-98040 |
| Progression disease, cfDNA levels | 25 | 11595.13 | 3747.6 | 18398.2 | 705.1-80440.3 |

**Supplementary Table 2.** Receiver operating characteristic curves analysis to determine the value of cfDNA levels to discriminate progression or death.

| Parameters | AUROC | Threshold | Sensitivity | Specificity |
|---|---|---|---|---|
| **PFS** | | | | |
| Log cfDNA at baseline | 0.834 | 7.650 | 0.854 | 0.800 |
| Log cfDNA at 3 weeks | 0.754 | 7.879 | 0.629 | 1.000 |
| **OS** | | | | |
| Log cfDNA at baseline | 0.783 | 8.077 | 0.789 | 0.750 |
| Log cfDNA at 3 weeks | 0.707 | 7.879 | 0.625 | 0.750 |
| Log cfDNA at progression disease | 0.606 | 8.495 | 0.545 | 1.000 |

**Supplementary Table 3.** Analysis to determine the prognostic value of CTCs to discriminate progression or death.

| **CellSearch® CTC levels** | | | | | | | |
|---|---|---|---|---|---|---|---|
| CTC, median (range) | 26 (0-4,796) | | | | | | |
| CTC, mean ± SE | 579.2 ± 284.5 | | | | | | |
| | | **PFS** | | | **OS** | | |
| **CellSearch® CTC count** | **n (%)** | **Median (days)** | **HR (95%CI)** | ***p*-value** | **Median (days)** | **HR (95%CI)** | ***p*-value** |
| 0 | 3 (14.29) | - | - | - | - | - | - |
| ≥1 | 18 (85.71) | 179 | 7.18 (0.92-56.3) | 0.07 | 236 | 4.98 (0.65-38.1) | 0.09 |
| ≥2 | 17 (80.95) | 179 | 3.93 (0.88-17.6) | 0.08 | 229 | 3.2 (0.72-14.2) | 0.08 |
| ≥5 | 13 (61.91) | 185 | 1.64 (0.61-4.41) | 0.4 | 244 | 1.62 (0.58-4.52) | 0.35 |
| ≥10 | 12 (57.14) | 179 | 1.72 (0.66-4.5) | 0.3 | 236 | 1.62 (0.6-4.41) | 0.2 |
| ≥50 | 9 (42.86) | 163 | 1.81 (0.7-4.67) | 0.11 | 229 | 1.71 (0.63-4.68) | 0.3 |
| ≥100 | 8 (38.1) | 171 | 1.63 (0.62-4.26) | 0.2 | 254 | 1.52 (0.55-4.25) | 0.5 |
| ≥150 | 6 (28.57) | 158 | 2.52 (0.81-7.86) | 0.03 | 217 | 2.32 (0.76-7.15) | 0.07 |
| ≥500 | 5 (23.81) | 163 | 2.8 (0.81-9.7) | 0.10 | 229 | 2.4 (0.73-7.9) | 0.2 |
| ≥1000 | 3 (14.29) | 154 | 4.6 (0.92-23) | 0.04 | 205 | 2.07 (0.46-9.43) | 0.2 |

Abbreviations: PFS, progression-free survival; OS, overall survival; HR, hazard ratio; CI, confidence interval.

**Supplementary Table 4**. Progression-free survival and overall survival probabilities estimated according two risk groups.

| **Risk Groups** | ***PFS*** | | | ***OS*** | | |
|---|---|---|---|---|---|---|
| | HR | 95% CI | p-value | HR | 95% CI | p-value |
| Favorable | 1 | (Ref.) | - | 1 | (Ref.) | - |
| Intermediate-poor | 5.37 | 2.32-12.4 | < 0.001 | 6.02 | 2.66-13.6 | < 0.001 |

Abbreviations: PFS, progression-free survival; OS, overall survival; HR, hazard ratio; CI, confidence interval.

**SUPPLEMENTARY FIGURES**

**Supplementary Figure 1.** CfDNA levels at different time-points (baseline, 3 weeks after treatment onset and at progression disease. Total cfDNA levels at baseline were significantly higher than at 3 weeks after the therapy onset (Wilcoxon test *p*= 0.002).

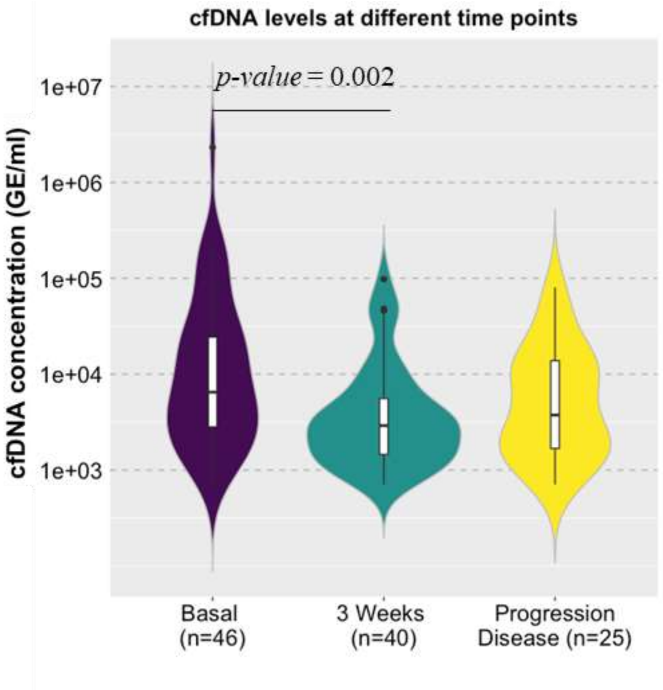

**Supplementary Figure 2.** (A) Example of CTCs detected in our cohort using the CellSearch® system. (B) Number of CTCs detected in each patient. Patients with presence of <10 CTCs are represented in red columns (C) The presence of ≥150 CTCs/7.5mL of blood was significantly associated with shorter PFS rates.

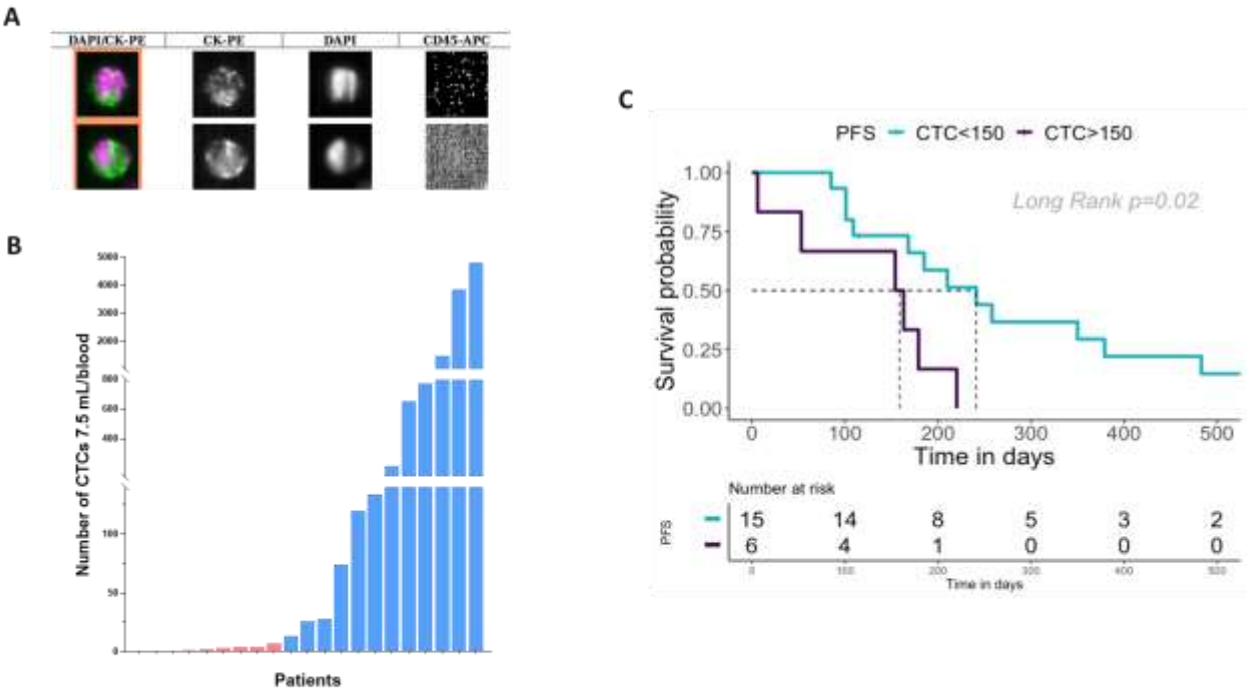

**Supplementary Figure 3.** CTCs and association with clinical characteristics. The number of CTCs detected in our cohort were significantly associated with the stage of patients and a poor performed status. Also, high cfDNA levels and the presence of CTCs at baseline were significantly associated.

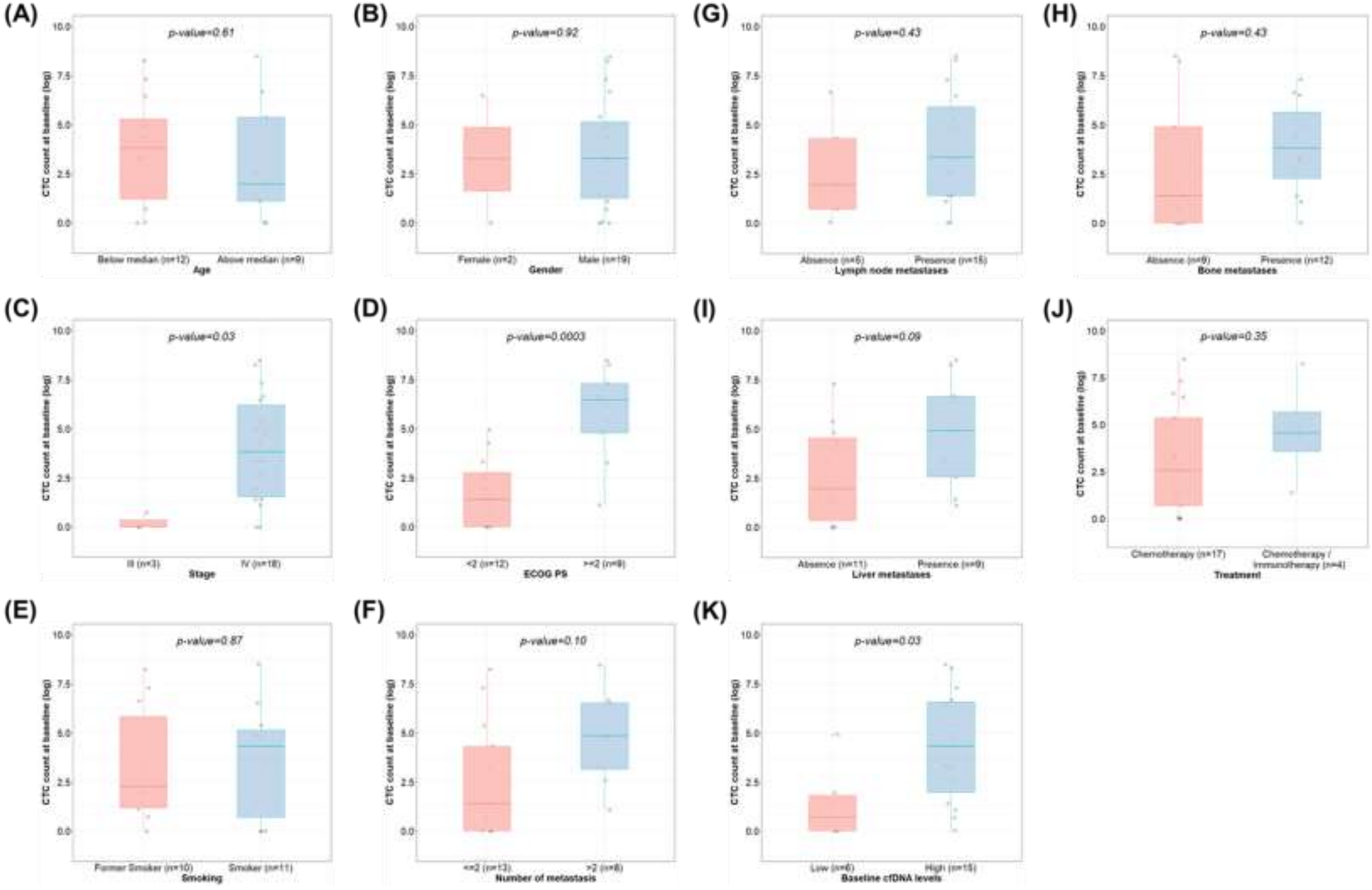
